# Assessment and Improvement of Elixhauser Comorbidity Index for Predicting In-hospital Mortality in Heart Transplant Patients

**DOI:** 10.1101/2023.03.27.23287837

**Authors:** Renxi Li

## Abstract

**Background:** Heart transplant (HT) has a high in-hospital mortality of around 5%. Risk prediction in-hospital mortality can be informative for transplant candidacy and post-HT prognosis. Elixhauser Comorbidity Index (ECI) is an ICD diagnostic code-based comorbidity measurement tool that can predict in-hospital mortality. While it has been validated in the large in-patient population, the accuracy of the mortality prediction has not been assessed in HT.

**Methods:** This study assessed the in-hospital mortality risk prediction by ECI as well as demographic variables in HT patients in the National Inpatient Sample (NIS) database. Demographic information was included in the multivariable ECI with demographics (ECID) model to assess in-hospital mortality. Moreover, ECI and age were used to develop a single index adjusted ECI (aECI) for mortality prediction.

**Results:** Age best predicts (*c*-statistic = 0.673, 95% CI = 0.638-0.709) in-hospital mortality, followed by ECI (*c*-statistic = 0.638, 95% CI = 0.598-0.678), race (*c*-statistic = 0.571, 95% CI = 0.533-0.609). Sex did not have predictive power (*c*-statistic = 0.501, 95% CI = 0.467-0.535) for in-hospital mortality. The predictive power of ECI was improved (*c*-statistic = 0.753, 95% CI = 0.720-0.785) in the ECID model. The single measure aECI had comparable discriminative power (*c*-statistic = 0.763, 95% CI = 0.731-0.794) to ECID in predicting in-hospital mortality.

**Conclusion:** This study showed that ECI was an effective measure to predict post-HT in-hospital mortality. The improved measure aECI can be easily derived from ECI as a quick reference to assess post-HT in-hospital mortality in both the clinic and health administration.

## Introduction

Heart transplant (HT) is an effective therapy to extend life for patients with advanced heart disease ^1^. As a high-risk procedure, the post-HT in-hospital period has a markedly high mortality of around 5% compared to 0.7% of all surgical procedures ^1–4^. Several risk factors, including age, race, renal failure, and liver function, have been identified ^1,3,4^. Thus, risk prediction post-HT in-hospital mortality is informative for transplant candidacy and post-HT prognosis ^4^.

Assessing clinical outcomes often needs to comorbidity for risk adjustment ^5,6^. The Elixhauser Comorbidity measure was developed to identify a comprehensive set of comorbidities in large-scale administrative data based on International Classification of Diseases, Ninth/Tenth Revision, Clinical Modification (ICD-9/10-CM) codes ^7^. There are 29 and 38 comorbidities identified in the ICD-9-CM and ICD-10-CM systems, respectively ^8^. Several studies have shown the effectiveness of using Elixhauser Comorbidity to identify the risk of in-hospital mortality ^9–11^. Recently, Elixhauser Comorbidity was further developed into Elixhauser Comorbidity Index (ECI), a weighted single scoring system, to predict in-hospital mortality ^12^.

ECI has shown overall high predictive power (*c*-statistic = 0.777, 95% CI = 0.776-0.778) for in-hospital mortality in all subjects during the validation phase of its development ^12^. While ECI has been validated in several disease categories with top mortality such as septicemia and pneumonia, in-hospital mortality in HT patients has not been specifically assessed by ECI ^12^.

This study aimed to assess the in-hospital mortality risk prediction by ECI in HT patients in the National (Nationwide) Inpatient Sample (NIS) database, the largest in-patient public database in the US that accounts for 20% of all hospital discharges ^13^. Since ECI does not include demographic information, in-hospital mortality risk predictions by demographics, including age, race, and sex, were also assessed. Finally, this study aimed to improve the predictive power of ECI by including relevant demographic information in the model to develop the single index – adjusted ECI (aECI).

## Materials and Methods

The NIS database was used to extract patients who received heart transplants using the International ICD-10 Procedure Coding System (ICD-10-PCS) by 02YA0Z0 between the last quarter of 2015 and 2020. Patient demographic information including age, race, and sex, in-hospital mortality record, and ICD-10-CM was extracted. Elixhauser Comorbidity Software was used to identify 38 Elixhauser Comorbidity and then calculate ECI ^8^. The weights for the Elixhauser Comorbidities are listed in Table 1.

**Table 1.** Elixhauser Comorbidities identified by ICD-10 Clinical Modification (ICD-10-CM) used to derive the Elixhauser Comorbidities Index (ECI) in 3,469 heart transplant patients from the last quarter of 2015 to 2020 in NIS.

| **Elixhauser Comorbidities** | **N** | **Percentage** | **Weight** |
| --- | --- | --- | --- |
| Acquired immune deficiency syndrome | 9 | 0.26% | -4 |
| Alcohol abuse | 44 | 1.27% | -1 |
| Anemias due to other nutritional deficiencies | 0 | 0.00% | -3 |
| Autoimmune conditions | 71 | 2.05% | 0 |
| Chronic blood loss anemia (iron deficiency) | 0 | 0.00% | -4 |
| Lymphoma | 21 | 0.61% | 9 |
| Leukemia | 12 | 0.35% | 5 |
| Metastatic cancer | 0 | 0.00% | 22 |
| Solid tumor without metastasis, in situ | 0 | 0.00% | 0 |
| Solid tumor without metastasis, malignant | 13 | 0.37% | 10 |
| Cerebrovascular disease | 110 | 3.17% | 5 |
| Heart failure | 645 | 18.59% | 14 |
| Coagulopathy | 0 | 0.00% | 14 |
| Dementia | 5 | 0.14% | 5 |
| Depression | 341 | 9.83% | -8 |
| Diabetes without chronic complications | 144 | 4.15% | -2 |
| Diabetes with chronic complications | 907 | 26.15% | 0 |
| Drug abuse | 54 | 1.56% | -7 |
| Hypertension, complicated | 1153 | 33.24% | 1 |
| Hypertension, uncomplicated | 293 | 8.45% | 0 |
| Liver disease, mild | 0 | 0.00% | 2 |
| Liver disease, moderate to severe | 4 | 0.12% | 16 |
| Chronic pulmonary disease | 403 | 11.62% | 2 |
| Neurological disorders affecting movement | 0 | 0.00% | -1 |
| Other neurological disorders | 0 | 0.00% | 22 |
| Seizures and epilepsy | 0 | 0.00% | 2 |
| Obesity | 413 | 11.91% | -7 |
| Paralysis | 79 | 2.28% | 4 |
| Peripheral vascular disease | 233 | 6.72% | 3 |
| Psychoses | 0 | 0.00% | -9 |
| Pulmonary circulation disease | 0 | 0.00% | 4 |
| Renal failure, moderate | 0 | 0.00% | 3 |
| Renal failure, severe | 32 | 0.92% | 7 |
| Hypothyroidism | 434 | 12.51% | -3 |
| Other thyroid disorders | 82 | 2.36% | -8 |
| Peptic ulcer disease x bleeding | 0 | 0.00% | 0 |
| Valvular disease | 230 | 6.63% | 0 |
| Weight loss | 0 | 0.00% | 13 |

Logistic regression was performed for a best-fit model to predict in-hospital mortality by ECI, age, race, and sex, respectively. Then, multivariable logistic regression was used to examine the best-fit prediction model for in-hospital mortality by the ECI with demographics (ECI-D) model, which included ECI, age, race, and sex. The receiver operating characteristic (ROC) for each regression model was plotted. The area under the ROC curve (AUC) was estimated with a 95% confidence interval for each model. The ability of each model to discriminate in-hospital mortality was characterized by *c*-statistic, where *c*-statistic = 0.5 represents the model has no predictive power and *c*-statistic = 1 means the model accurately classifies all in-hospital death. Generally, *c*-statistic > 0.7 indicates the presence of good predictive variables.

Since race and sex had very small predictive power for in-hospital mortality, ECI and age were chosen to develop the single index aECI based on Sullivan’s method ^14,15^. The parameter estimates of ECI and age from the ECID multivariable regression were extracted. Weights should be calculated by dividing all regression coefficients by the smallest regression coefficient and then rounding the quotient to the nearest integer. This way, a larger weight indicates a stronger association by proportion to the variable with the smallest coefficient. In this case, however, there were only two variables (i.e. ECI and age) and the quotient of their regression coefficients was around 1.5; rounding to the nearest integer would introduce too much error. So instead, weights were calculated by dividing the regression coefficients by half of the smallest coefficient, followed by rounding the quotient to the nearest integer. A weighted addition of ECI and age was used to calculate a single index of aECI.

All analyses were performed by SAS version 9.4. The study was retrospective based on open database and was exempted from IRB approval.

## Results

There were a total number of 3,469 HT patients identified between the last quarter of 2015 and 2020 from NIS. In the cohort, Elixhauser Comorbidity is summarized in Table 1, where 1,153 complicated hypertension (33.24%), 907 diabetes with chronic complications (26.15%), 645 heart failure (18.59%), 434 hypothyroidism (12.51%), 413 obesity (11.91%), 403 chronic pulmonary disease (11.62%), 341 depression (9.83%), 293 uncomplicated hypertension (8.45%), 233 peripheral vascular disease (6.72%), 230 valvular disease (6.63%), 144 diabetes without chronic complications (4.15%), 110 cerebrovascular disease (3.17%), 82 other thyroid disorders (2.36%), 79 paralysis (2.28%), 71 autoimmune conditions (2.05%), 54 drug abuse (1.56%), 44 alcohol abuse (1.27%), 32 severe renal failure (0.92%), 21 lymphoma (0.61%),13 malignant solid tumor without metastasis (0.37%), 12 leukemia (0.35%), 9 acquired immune deficiency syndrome (0.26%), 5 dementia (0.14%), and 4 moderate to severe liver disease, (0.12%) were identified. No patient with anemias due to other nutritional deficiencies, chronic blood loss anemia (iron deficiency), metastatic cancer, in situ solid tumor without metastasis, coagulopathy, mild liver disease, neurological disorders affecting movement, seizures and epilepsy, psychoses, pulmonary circulation disease, moderate renal failure, peptic ulcer disease x bleeding, weight loss, or other neurological disorders was identified.

The demographics and characteristics of patients were summarized in Table 2. The mean age of the cohort was 47.79 ± 19.79 years old. HT recipient age showed an overall left-skewed distribution and a spike in infant heart transplantation. The racial group was composed of 1983 (57.16%) white, 711 (20.50%) African Americans, 362 (10.44 %) Hispanics, 132 (3.81 %) Asians, 14 (0.40 %) American Indians, and 113 (3.26 %) other races. There were 2462 (70.97%) males and 1007 (29.03%) females identified. A total number of 178 in-hospital mortality were recorded with an overall mortality rate of 5.13%. The mean ECI was 0.97 ± 7.04 with a minimal score of -30 and a maximal score of 25. The distribution of age and ECI were visualized in Figure 1, which showed a roughly normal distribution.

**Table 2.**
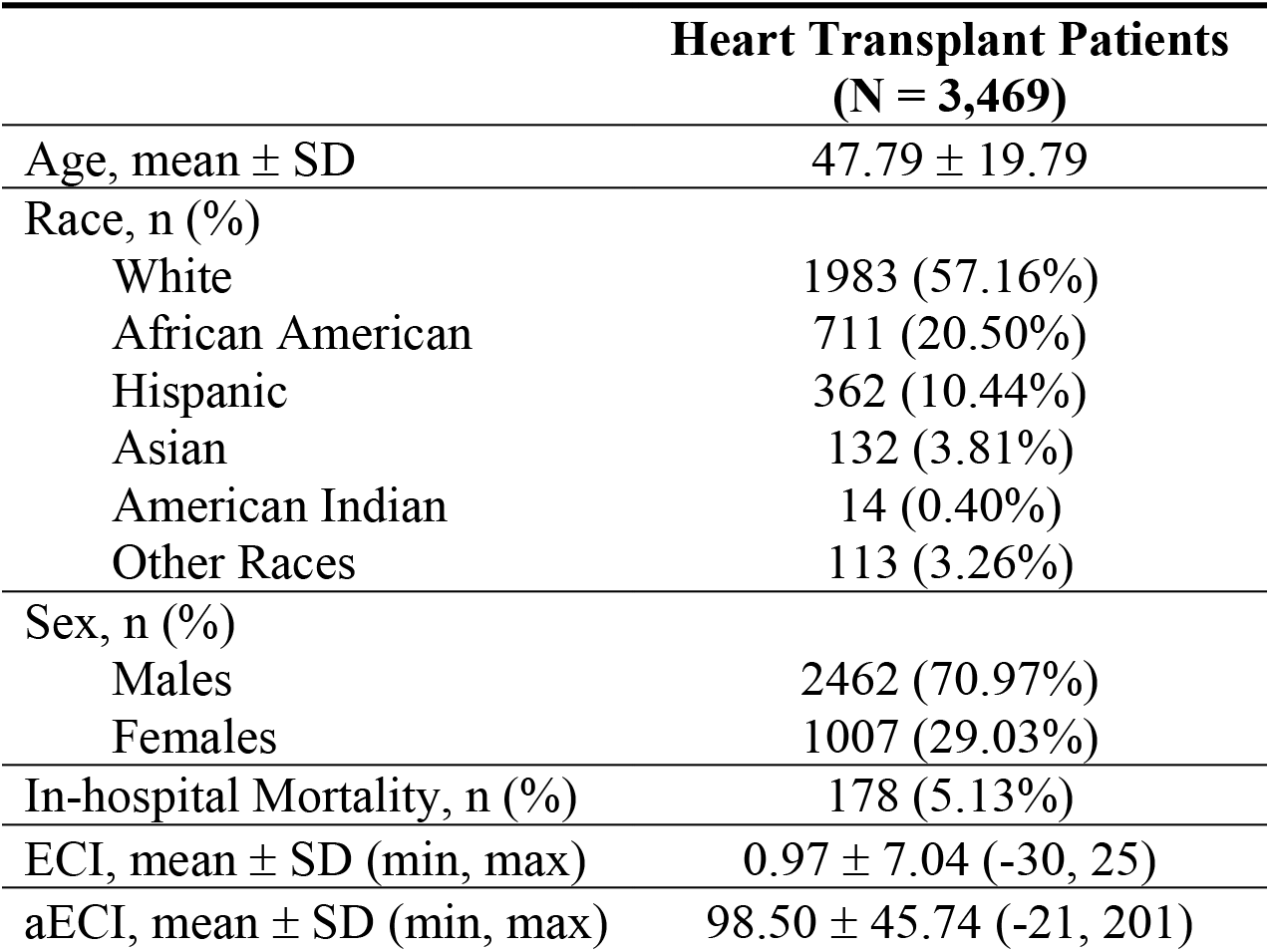
Demographics and characteristics of heart transplant patients from the last quarter of 2015 to 2020 identified in NIS. ***Abbreviations:*** aECI, adjusted Elixhauser Comorbidity Index; ECI, Elixhauser Comorbidities Index; SD, standard deviation.

**Figure 1.**
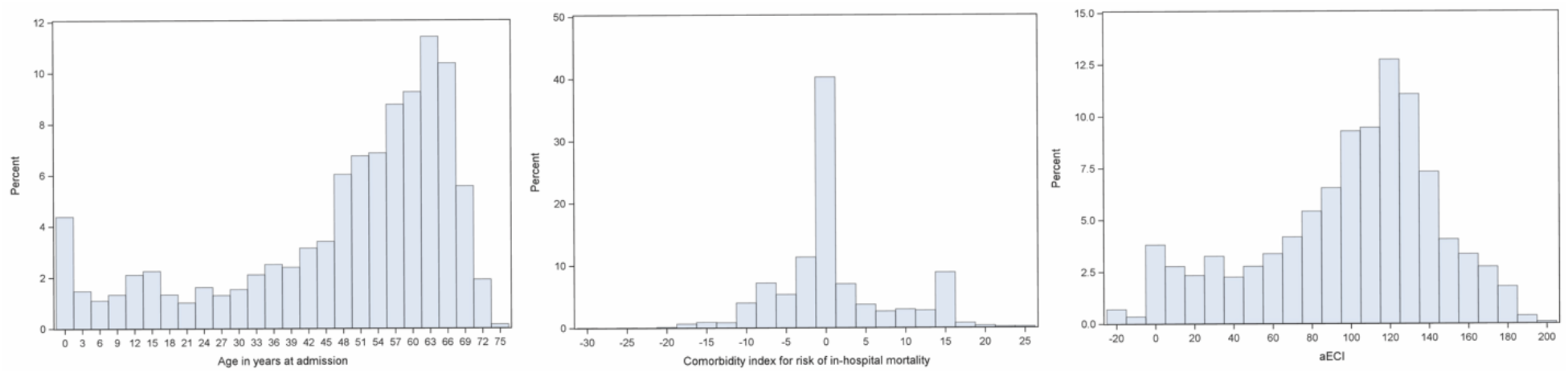
Distribution of age and Elixhauser Comorbidity Index (ECI) of patients who underwent heart transplants from the last quarter of 2015 to 2020 identified in NIS.

A summary of the *c*-statistic for ECI, age, race, and sex was shown in Figure 2 and summarized in Table 3. Age best predicts (*c*-statistic = 0.673, 95% CI = 0.638-0.709) in-hospital mortality, followed by ECI (*c*-statistic = 0.638, 95% CI = 0.598-0.678), race (*c*-statistic = 0.571, 95% CI = 0.533-0.609). Sex did not have predictive power (*c*-statistic = 0.501, 95% CI = 0.467-0.535) for in-hospital mortality. None of the single parameters had a significant *c*-statistic (over 0.70). By including ECI, age, race, and sex in one multivariable logistic regression, the result for the ECID model was shown in Figure 3 and summarized in Table 3. The ECID model had significant discriminative power (*c*-statistic = 0.753, 95% CI = 0.720-0.785) for in-hospital mortality. The results for the aECI model were shown in Figures 1 and 3 and summarized in Tables 2 and 3. The weights for ECI and age were 3 and 2, respectively, so aECI is calculated as aECI = 2*age+3*ECI. The mean aECI was 98.50 ± 45.74 with a minimal score of -21 and a maximal score of 201. The distribution of aECI was shown in Figure 1, showing a roughly normal distribution. The single index aECI had comparable predictive power (*c*-statistic = 0.763, 95% CI = 0.731-0.794) for in-hospital mortality compared to the multivariable ECID model.

**Figure 2.**
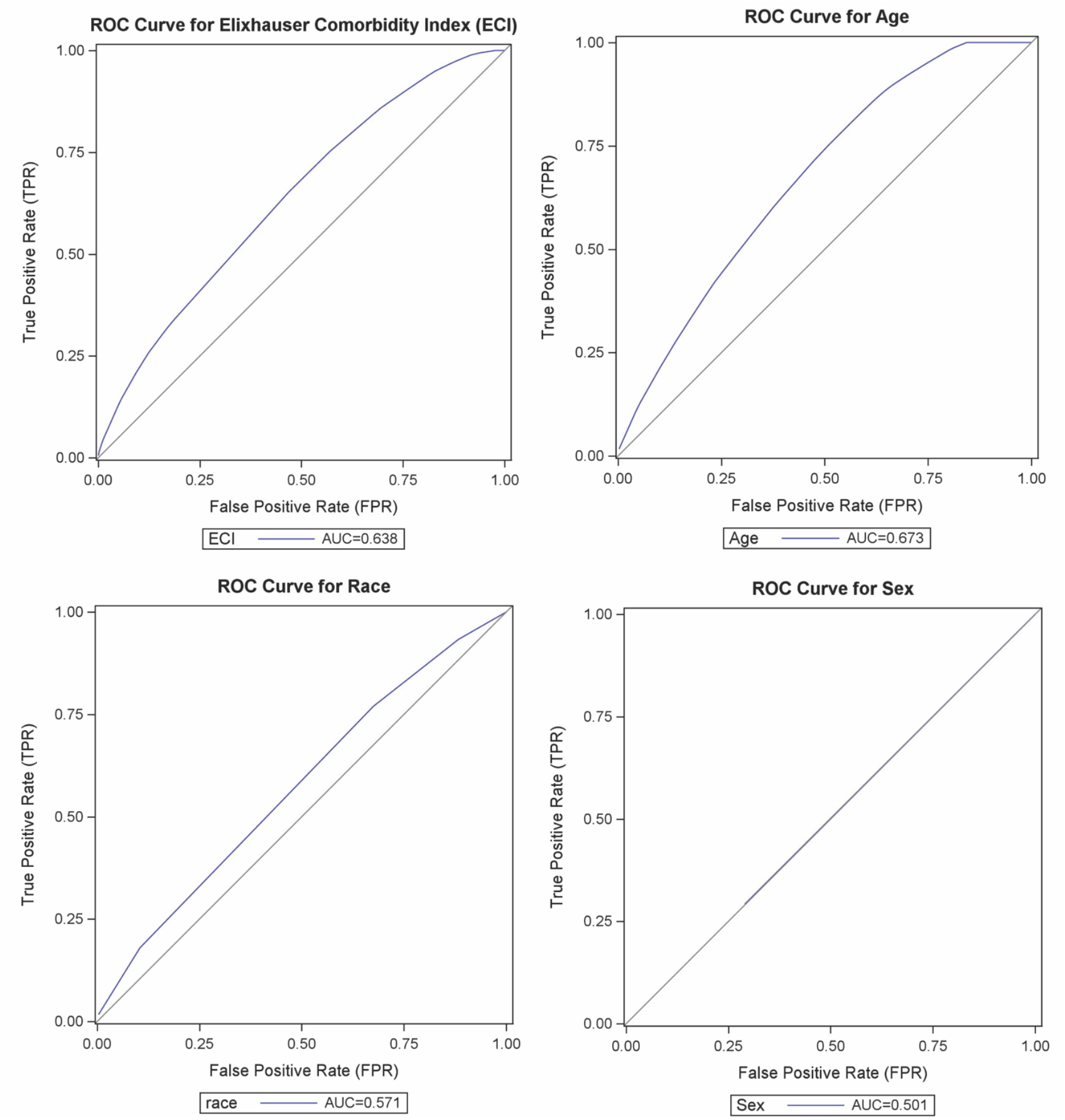
Receiver operating characteristic (ROC) curves for logistic regression models predicting in-hospital mortality by ECI, age, race, or sex, respectively, in patients who underwent heart transplants from the last quarter of 2015 to 2020 in NIS. A straight diagonal line shows the null model (AUC 0.500).

**Table 3.**
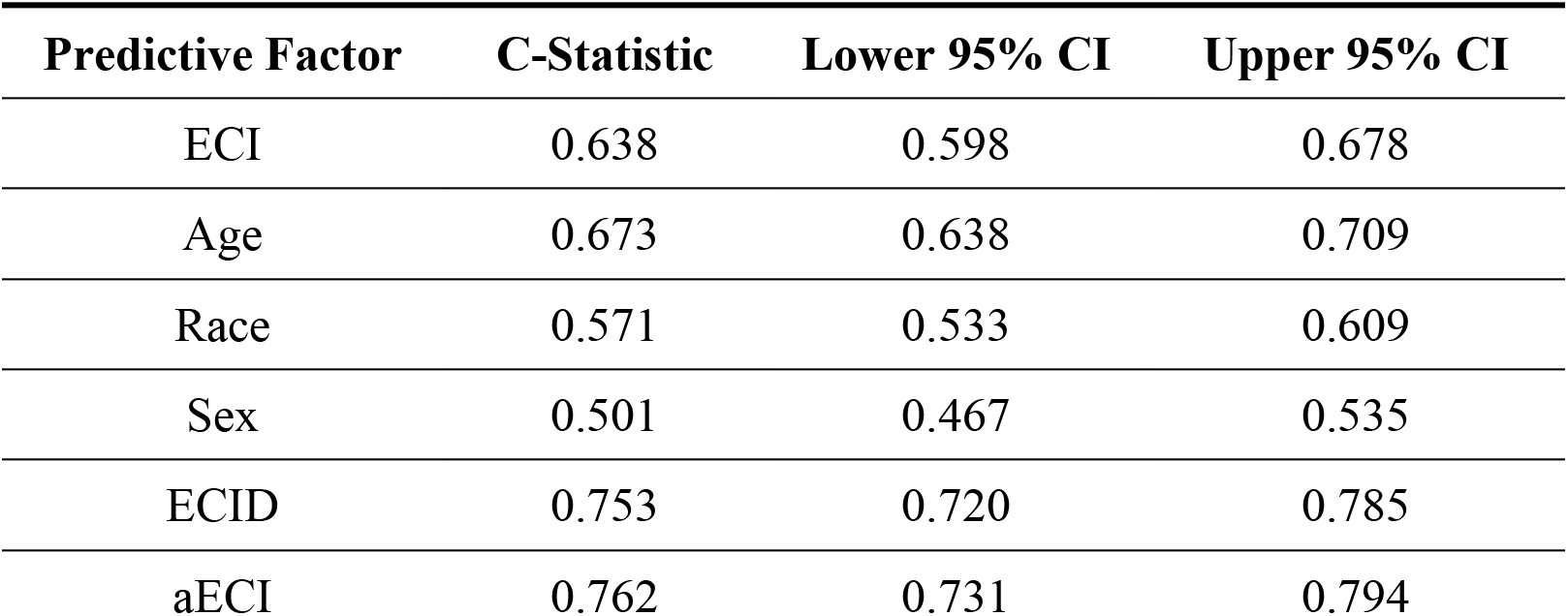
Discrimination for in-hospital mortality by predictor factors in heart transplant patients from the last quarter of 2015 to 2020 in NIS. The ECID model includes ECI, age, race, and sex. aECI is a single index that is calculated by 2*age+3*ECI. ***Abbreviations:*** aECI, adjusted Elixhauser Comorbidity Index; CI, confidence interval; ECI, Elixhauser Comorbidities Index; ECID, Elixhauser Comorbidity Index with Demographics.

| Predictive Factor | C-Statistic | Lower 95% CI | Upper 95% CI |
| --- | --- | --- | --- |
| ECI | 0.638 | 0.598 | 0.678 |
| Age | 0.673 | 0.638 | 0.709 |
| Race | 0.571 | 0.533 | 0.609 |
| Sex | 0.501 | 0.467 | 0.535 |
| ECID | 0.753 | 0.720 | 0.785 |
| aECI | 0.762 | 0.731 | 0.794 |

**Figure 3.**
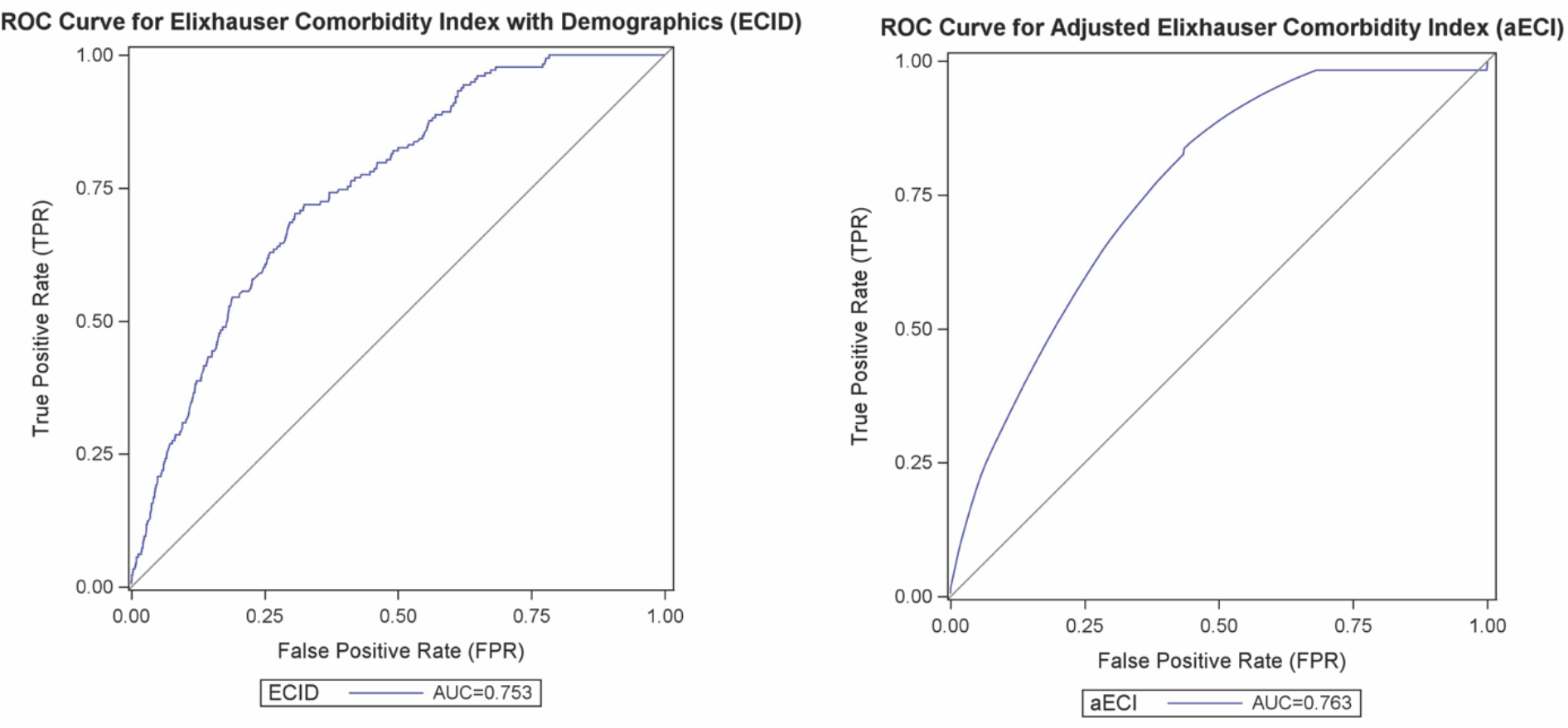
Receiver operating characteristic (ROC) curves for predicting in-hospital mortality by multivariable logistic regression of ECID model and logistic regression of aECI respectively, in patients who underwent heart transplants from 2015 to 2020 in NIS. A straight diagonal line shows the null model (AUC 0.500). The ECID model includes ECI, age, race, and sex. aECI is a single index that is calculated by 2*age+3*ECI.

## Discussion

This study examined predicted mortality by ECI and demographics in recent HT patients in NIS data from the last quarter of 2015 to 2020. ECI and Age were found to be moderate predictors for in-hospital mortality while race and sex were weaker predictors. A multivariable regression including ECI and all three demographic variables increased the predictor model to a significant level. ECI and age were chosen to develop a new single index aECI, which showed comparable effectiveness in predicting in-hospital mortality in HT.

ECI gave a moderate but not significant prediction (*c*-statistic = 0.638) of in-hospital mortality post-HT. Thus, comorbidities alone did not have sufficient discriminative power and the inclusion of additional demographic variables was justified.

Recipient age at transplantation was the strongest single predictor for in-hospital mortality in HT patients. This was incongruent with previous studies, where Singh et al identified age > 65 as a risk factor (odds ratio = 1.89) in their risk stratification model ^4^. It was proposed that aging compromised the immune function which lead to an increase in all-cause mortality ^16^.

Race had little effect on post-HT in-hospital mortality, which confirmed the previous results from Singh et al where all races benefited equally from HT ^3^. However, Singh et al pointed out that long-term survival only improved in Causation but not African Americans or Hispanics ^3^. Additional racial modifiers might need to be included in aECI formula if future studies explore the long-term prediction of mortality by ECI.

Sex did not affect the prediction of in-hospital mortality in HT, which was consistent with previous studies ^17,18^. Sex-mismatch (e.g. female heart in male recipient), however, has been widely shown to have a worse prognosis ^19–22^. The effect of sex-mismatch can be examined in future studies if donor information is recorded.

With additional demographic information included, the multivariable ECID offered a much better prediction of mortality. The further simplification of ECID with only ECI and age into a single aECI score maintained the high predictive power. aECI (*c*-statistic = 0.763) could better discriminate the presence of mortality than the Singh model (*c*-statistic = 7.22), where age, specific diagnosis, mechanical support type, ventilator support, estimated glomerular filtration rate, and total serum bilirubin were used in the model ^4^.

ECI calculations are free and readily accessible. For clinicians and patients, ECI can be calculated by a free online ECI calculator based on past medical history ^23^. For researchers and healthcare administrators, ECI can be calculated by the Elixhauser Comorbidity Software from NIS ^8^. Given that aECI can be easily derived from ECI, aECI has the advantage of being a quick reference for post-HT in-hospital mortality without the need for lab tests.

There were several limitations of this study. First, all mortality examined was in-hospital without any follow-up. It was argued that to examine early post-transplant outcomes, a 30-day perioperative period should be followed ^24^. Second, Elixhauser Comorbidities were recorded as binary (present/not present) so that within the same comorbidity, patients were more heterogenous with different extents/staging of the disease. Also, no specific diagnosis, pertinent medical history, or lab values associated with the cardiovascular diseases were included, which means aECI under-appreciate the clinical complexation of the patients. Thus, aECI should be used in conjunction with other clinical information to assess the mortality risk. Lastly, relevant donor factors, such as donor age, sex, and ischemic time, are not included in the model.

Nevertheless, this study showed the effectiveness of ECI and aECI in predicting post-HT in-hospital mortality. Future studies can examine the long-term predictor power of ECI in post-HT mortality, as well as include clinical data and/or donor information in the prediction model.

## Data Availability

All data produced in the present study are available upon reasonable request to the authors

## Abbreviations

aECI: adjusted Elixhauser Comorbidity Index;
AUC: area under curve;
CI: confidence interval;
ECI: Elixhauser Comorbidity Index;
ECI-D: Elixhauser Comorbidity Index with demographics;
HT: Heart transplant;
ICD-9/10-CM: International Classification of Diseases, Ninth/Tenth Revision, Clinical Modification;
NIS: National (Nationwide) Inpatient Sample;
ROC: receiver operating characteristic.

## Notes

**Disclosure:** The author declares no conflicts of interest.

### Competing Interest Statement

The authors have declared no competing interest.

### Funding Statement

This study did not receive any funding

### Author Declarations

The study used only openly available human data from The National (Nationwide) Inpatient Sample (NIS) that were originally located at https://hcup-us.ahrq.gov/databases.jsp

